# Survival outcomes of rectal and head and neck cancer patients receiving radio-(chemo)therapy with a ketogenic diet. Results from a controlled clinical study (KETOCOMP)

**DOI:** 10.1101/2025.07.23.25332053

**Authors:** Rainer J. Klement, Reinhart A. Sweeney

## Abstract

**Background:** High-fat, low carbohydrate ketogenic diets (KDs) have been proposed to target the cancer hallmark of high glycolytic metabolism and synergize with radio- and chemotherapy. We here report survival outcomes of rectal and head and neck cancer (HNC) patients who underwent a KD during radio-(chemo)therapy.

**Materials and Methods:** Patients had participated in the controlled KETOCOMP study. The intervention group had consumed a KD during radiotherapy, while the control group had maintained their standard diet. All patients were prospectively followed until the occurrence of disease progression or death to evaluate overall (OS), progression-free (PFS) and locoregional recurrence-free survival (RFS) with the Kaplan-Meier method and by computing restricted mean survival times. In order to simulate a randomized controlled trial and meta-analysis, patients in the KD groups were matched to control patients with propensity score matching and data was pooled.

**Results:** Median follow-up was 77.4 (range 12.1-107.9) months (HNC) and 72.6 (16.5-127.1) months (rectal cancer), respectively. HNC patients exhibited non-significantly longer OS and PFS and a trend for longer RFS times (p=0.079). These trends became more pronounced in the propensity score-matched sample (log-rank test p-values <0.10 for all three outcomes). In rectal cancer patients, there was no evidence for a benefit of the KD group. Analysis of the matched and pooled sample revealed a significantly longer restricted mean PFS time in the KD group (p=0.027) and a trend for longer RFS times (p=0.062). OS was also longer in the pooled KD group, but not significantly (p=0.29).

**Conclusions:** These data indicate the potential for synergistic effects of a KD combined with radio-(chemo)therapy. While sample sizes could have prohibited the observed survival differences from becoming statistically significant, our data are useful to inform future studies or meta-analyses.

## Introduction

For more than 100 years, an altered metabolism has been recognized as a defining hallmark of cancer. In the 1920s, Otto Warburg and his co-workers showed that, compared to normal tissues, tumor tissue consumes large amount of glucose and in turn excretes large amounts of lactate, a phenomenon now denoted as the Warburg effect [1]. Meanwhile, data have been obtained indicating that cancer cell mitochondria are frequently dysmorphic and dysfunctional, which would explain the compensatory upregulation of glucose fermentation to lactate and other fermentative pathways such as glutaminolysis to maintain the cancer cell’s necessary energy yield [2,3]. In addition, cancer cells utilize glucose in the pentose phosphate pathway (PPP) to promote their proliferation. The PPP typically yields ribose for DNA and RNA synthesis, but could also yield lactate plus acetyl-CoA for fatty acid synthesis in cancer cells with a mutated transketolase-like 1 (TKTL1) enzyme [4].

Roughly 20 years after Otto Warburg’s seminal studies, a German oncologist named Wilhelm Brünings conducted clinical trials in which he attempted to target the Warburg effect by cutting tumors from their glucose supply using a ketogenic diet (KD) in combination with maximally tolerable insulin injections [5]. This was the first clinical application of ketogenic metabolic therapy (KMT). Today, ketogenic metabolic therapy utilizes metabolic inhibitors, KDs, ketogenic supplements and other applications targeting the weaknesses that emerge due to cancer cells’ altered metabolism and the accompanied metabolic inflexibility [6]. KDs are a central element of this therapeutic approach. Although preclinical studies have shown promising results of applying KDs to treat various cancer entities [7,8], clinical data with hard clinical endpoints are still sparse.

In 2016, we had initiated a controlled clinical trial to test the effects of a KD consumed during radiotherapy in patients with breast, head and neck or rectal cancer, respectively [9–12]. While the main aim was to test the effects of the KD on body composition changes during radiotherapy, a secondary goal was to study the putative anti-tumor efficacy by prospectively following patients after therapy completion. This paper reports the final results concerning clinical outcomes of patients with head and neck cancer (HNC) or rectal cancer who had received radiotherapy or radiochemotherapy, respectively, combined with a KD.

## Materials and Methods

The KETOCOMP study was a monocentric non-randomized controlled clinical trial performed at the Department of Radiotherapy and Radiation Oncology at the Leopoldina Hospital Schweinfurt, Germany [9]. Ethical approval was granted by the ethics committee of the Bavarian Medical Association (Landesaerztekammer Bayern), and the protocol was registered on August 6th 2015 under ClinicalTrials.gov identifier NCT02516501. In principal, patients with breast cancer, HNC or rectal cancer referred to our clinic for radiotherapy were eligible to participate. Summarized briefly, the recruitment was performed in two blocks. The first block of patients was enrolled into a control group consuming their usual standard diet (SD). The second block of patients was enrolled into the intervention group consisting of the switch to a KD (so as to minimize intergroup influencing in the waiting rooms/etc.). Patients who wanted to eat a KD were enrolled into the KD arm irrespective of the planned allocation to control or intervention arm, while patients not willing to consume a KD were offered the option to enroll into the SD group instead (see for more details). The primary outcomes of interest were differences between both arms with respect to body composition changes assessed by weekly bioelectrical impedance analysis. Secondary outcomes were changes in several quality of life metrics that we have already published [13,14]. Another secondary aim was to prospectively follow patients for at least five years or until the occurrence of locoregional or distant progression. This allows us to test for any putative differences between both arms with respect to overall survival (OS), progression-free survival (PFS) and locoregional relapse-free survival (RFS). We here report the results of survival analyses for the HNC and rectal cancer patient cohorts.

### Patients

Details of patient enrollment and characteristics have been published previously [11,12]. Briefly, 32 patients with HNC and 49 patients with rectal cancer all receiving neoadjuvant or adjuvant radio-(chemo-)therapy, respectively, were included in the study. Of those, 11 HNC and 24 rectal cancer patients were allocated to the KD group. One patient with rectal cancer who had followed a KD developed a fatal Fournier’s gangrene and died a few days after completing radiotherapy due to septic shock [15]; because his death was unrelated to his cancer or therapy, this patient was removed from all statistical analyses.

After treatment, follow-up care of every patient was performed regularly. Briefly, the patients were admitted to our clinic for follow-up care a few weeks after therapy and then yearly. Should they live too far away, regular follow-up letters were requested from the cooperating specialist clinics, where the patients continued to receive follow-up care. During April and May 2025, a final update of the study database was performed with an attempt to gather information from every patient from within the last year.

### Statistical analyses

The primary outcome of interest for this study was OS and PFS, both calculated according to the Kaplan–Meier method. OS was defined as the time since the start of radiotherapy until death from any cause. PFS was defined as the time since the start of radiotherapy until locoregional or distant tumor progression. RFS was defined as the time since the start of radiotherapy until locoregional disease progression. In case of no event, time-to-event data were censored at the last follow-up time. We also computed restricted mean survival times (RMSTs) in both the KD and SD groups that could be useful for future meta-analyses [16]. The RMST is a measure of average survival up to a specified follow-up time, which distinguishes it from the mean survival time *µ*:

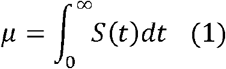

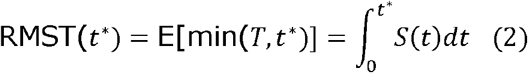

The RMST measures the area under the survival curve up to a specified truncation time. The truncation time *t*^*^ is given as

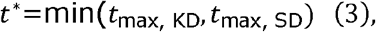

where *t*_max, KD_ and *t*_max, SD_ are the maximum follow-up times in the KD and SD groups for the outcome of interest (i.e., OS, PFS or RFS).

Survival analyses were performed for both the intention-to-treat (ITT) and per protocol (PP) populations. The ITT population was comprised of all patients starting the study and completing radiotherapy. The PP population is the subset of the ITT population ending the study without violating the protocol or voluntarily deciding to drop out. We also conducted survival analysis on a combined dataset of propensity-score-matched patients from the PP populations, in this way simulating evidence amalgamation from two randomized controlled trials. Propensity score matching was performed by submitting the two data sets to a logistic regression analysis, predicting the intervention group “KD” vs. “SD” with the set of variables {age, BMI, chemotherapy} for HNC patients and {age, BMI, PTV} for rectal cancer patients (PTV denotes the planning target volume). For each patient from the KD group one patient from the SD group was selected based on genetic matching, which is a robust method for achieving covariate balance [17].

Qualitative data are described using frequency and percentage distributions and compared between groups using Fisher’s exact test. Quantitative data are described using their median and range or mean and standard deviation and compared between groups using the Wilcoxon rank sum test or t-test, respectively. Kaplan-Meier survival curves were compared using the log rank test. Statistical significance was defined as p-values <0.05.

All analyses were performed in R version 4.4.1 with the survival and survminer packages for survival analysis, the survRM2 package for computing RMSTs and the MatchIt package for propensity score matching, thereby setting the pop.size parameter to 1000 [18]; for genetic matching, the MatchIt package calls functions from the Matching package [19].

## Results

### Patient characteristics

Table 1 summarizes the baseline characteristics of the ITT population. There were no statistically significant differences between the KD and SD groups with the exception that rectal cancer patients in the KD group had significantly smaller planning target volumes than rectal cancer patients in the control group (p=0.0018). When only the PP population was considered, this difference in PTV sizes remained statistically significant (p=0.0045). while all other differences between KD and SD arms remained nominally non-significant (all p≥0.05; see Ref. [11] and [12]).

**Table 1:**
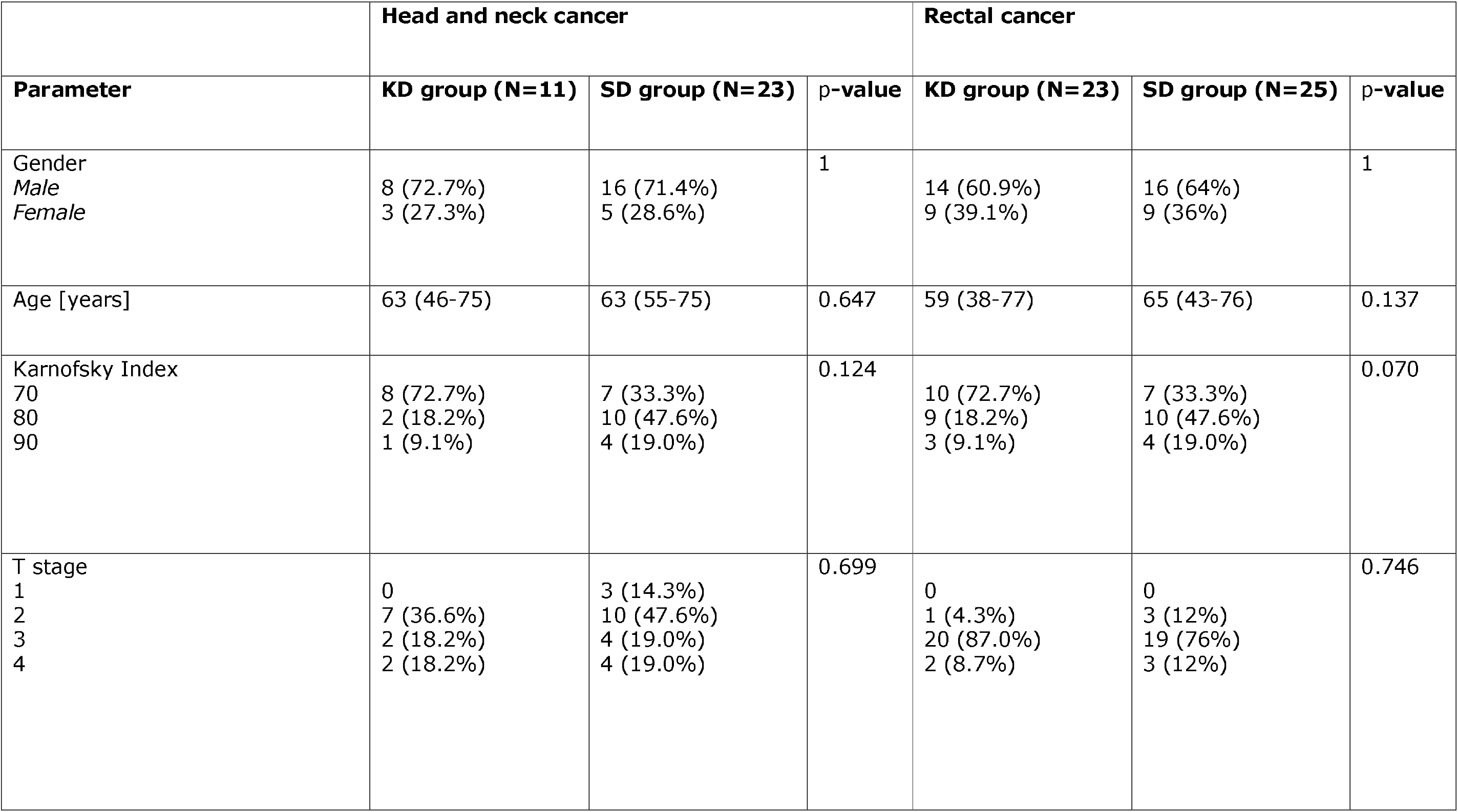
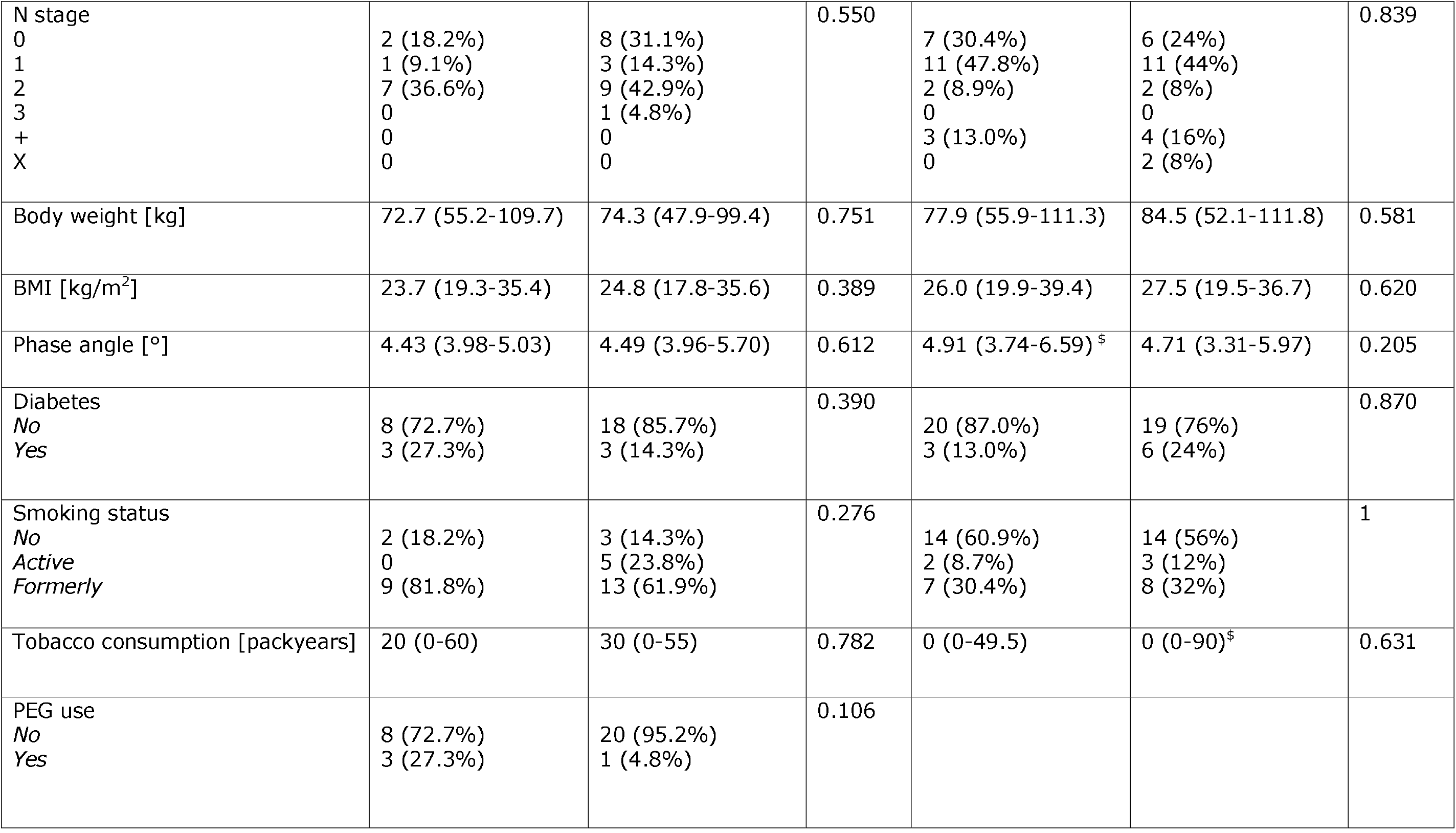
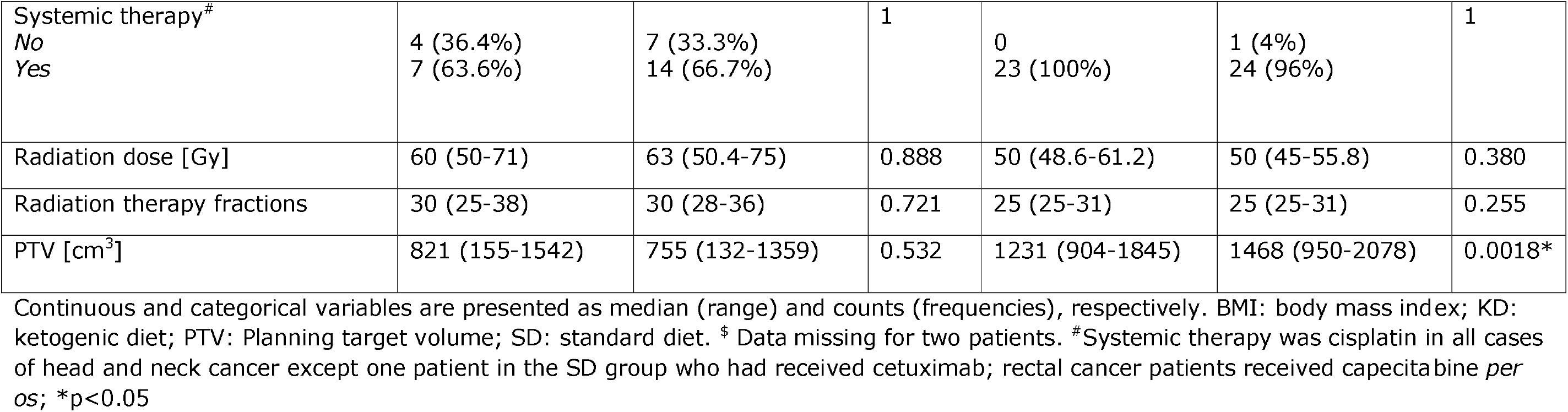
Baseline characteristics of the intervention and control groups.

|  | Head and neck cancer |  |  | Rectal cancer |  |  |
| --- | --- | --- | --- | --- | --- | --- |
| Parameter | KD group (N=11) | SD group (N=23) | p-value | KD group (N=23) | SD group (N=25) | p-value |
| Gender<br>*Male*<br>*Female* | 8 (72.7%)<br>3 (27.3%) | 16 (71.4%)<br>5 (28.6%) | 1 | 14 (60.9%)<br>9 (39.1%) | 16 (64%)<br>9 (36%) | 1 |
| Age [years] | 63 (46-75) | 63 (55-75) | 0.647 | 59 (38-77) | 65 (43-76) | 0.137 |
| Karnofsky Index<br>70<br>80<br>90 | 8 (72.7%)<br>2 (18.2%)<br>1 (9.1%) | 7 (33.3%)<br>10 (47.6%)<br>4 (19.0%) | 0.124 | 10 (72.7%)<br>9 (18.2%)<br>3 (9.1%) | 7 (33.3%)<br>10 (47.6%)<br>4 (19.0%) | 0.070 |
| T stage<br>1<br>2<br>3<br>4 | 0<br>7 (36.6%)<br>2 (18.2%)<br>2 (18.2%) | 3 (14.3%)<br>10 (47.6%)<br>4 (19.0%)<br>4 (19.0%) | 0.699 | 0<br>1 (4.3%)<br>20 (87.0%)<br>2 (8.7%) | 0<br>3 (12%)<br>19 (76%)<br>3 (12%) | 0.746 |
| N stage<br>0<br>1<br>2<br>3<br>+<br>X | 2 (18.2%)<br>1 (9.1%)<br>7 (36.6%)<br>0<br>0<br>0 | 8 (31.1%)<br>3 (14.3%)<br>9 (42.9%)<br>1 (4.8%)<br>0<br>0 | 0.550 | 7 (30.4%)<br>11 (47.8%)<br>2 (8.9%)<br>0<br>3 (13.0%)<br>0 | 6 (24%)<br>11 (44%)<br>2 (8%)<br>0<br>4 (16%)<br>2 (8%) | 0.839 |
| Body weight [kg] | 72.7 (55.2-109.7) | 74.3 (47.9-99.4) | 0.751 | 77.9 (55.9-111.3) | 84.5 (52.1-111.8) | 0.581 |
| BMI [kg/m<sup>2</sup>] | 23.7 (19.3-35.4) | 24.8 (17.8-35.6) | 0.389 | 26.0 (19.9-39.4) | 27.5 (19.5-36.7) | 0.620 |
| Phase angle [°] | 4.43 (3.98-5.03) | 4.49 (3.96-5.70) | 0.612 | 4.91 (3.74-6.59) <sup>§</sup> | 4.71 (3.31-5.97) | 0.205 |
| Diabetes<br>*No*<br>*Yes* | 8 (72.7%)<br>3 (27.3%) | 18 (85.7%)<br>3 (14.3%) | 0.390 | 20 (87.0%)<br>3 (13.0%) | 19 (76%)<br>6 (24%) | 0.870 |
| Smoking status<br>*No*<br>*Active*<br>*Formerly* | 2 (18.2%)<br>0<br>9 (81.8%) | 3 (14.3%)<br>5 (23.8%)<br>13 (61.9%) | 0.276 | 14 (60.9%)<br>2 (8.7%)<br>7 (30.4%) | 14 (56%)<br>3 (12%)<br>8 (32%) | 1 |
| Tobacco consumption [packyears] | 20 (0-60) | 30 (0-55) | 0.782 | 0 (0-49.5) | 0 (0-90) <sup>§</sup> | 0.631 |
| PEG use<br>*No*<br>*Yes* | 8 (72.7%)<br>3 (27.3%) | 20 (95.2%)<br>1 (4.8%) | 0.106 |  |  |  |
| Systemic therapy<sup>#</sup><br>No<br>Yes | 4 (36.4%)<br>7 (63.6%) | 7 (33.3%)<br>14 (66.7%) | 1 | 0<br>23 (100%) | 1 (4%)<br>24 (96%) | 1 |
| Radiation dose [Gy] | 60 (50-71) | 63 (50.4-75) | 0.888 | 50 (48.6-61.2) | 50 (45-55.8) | 0.380 |
| Radiation therapy fractions | 30 (25-38) | 30 (28-36) | 0.721 | 25 (25-31) | 25 (25-31) | 0.255 |
| PTV [cm<sup>3</sup>] | 821 (155-1542) | 755 (132-1359) | 0.532 | 1231 (904-1845) | 1468 (950-2078) | 0.0018* |

Continuous and categorical variables are presented as median (range) and counts (frequencies), respectively. BMI: body mass index; KD: ketogenic diet; PTV: Planning target volume; SD: standard diet. <sup>\$</sup> Data missing for two patients. <sup>#</sup>Systemic therapy was cisplatin in all cases of head and neck cancer except one patient in the SD group who had received cetuximab; rectal cancer patients received capecitabine *per os*; \*p<0.05

### Survival analysis for head and neck cancer patients

Table 2 shows the survival statistics for the HNC cohort. The length of follow-up was similar between the KD and SD groups for all three outcomes in both the ITT and PP population. The KD group exhibited longer RMSTs than the SD group, although the differences were not statistically significant. There were three locoregional relapses in the SD (14%), but none in the KD group, which resulted in a trend for an improved RMST in the KD group.

**Table 2:** Survival statistics of the intention-to-treat (ITT) and per protocol (PP) populations of head and neck cancer patients.

|  | Intention-to-treat population |  |  | Per protocol population |  |  |
| --- | --- | --- | --- | --- | --- | --- |
| Parameter | KD group (N=11) | SD group (N=23) | p-value | KD group (N=7) | SD group (N=25) | p-value |
| OS: Follow-up time [months] | 72.2 (12.4-102.2) | 88.2 (12.1-107.9) | 0.242 | 80.4 (12.4-102.2) | 88.2 (12.1-107.9) | 0.811 |
| OS: RMST [months] | 86.08±10.31 | 77.75±7.24 | 0.508 | 89.37±11.88 | 77.74±7.24 | 0.403 |
| OS: RMST ratio | 1.11±0.17 |  | 0.502 | 1.15±0.19 |  | 0.391 |
| PFS: Follow-up time [months] | 61.4 (4.3-100.5) | 77.1 (11.1-101.1) | 0.271 | 77.3 (4.3-100.5) | 77.1 (11.1-101.1) | 0.678 |
| PFS: RMST [months] | 83.96±10.60 | 73.23±7.69 | 0.413 | 86.76±12.72 | 73.23±7.69 | 0.363 |
| PFS: RMST ratio | 1.15±0.19 |  | 0.405 | 1.19±0.22 |  | 0.347 |
| RFS: Follow-up time [months] | 61.4 (9.9-100.5) | 74.4 (6.2-101.1) | 0.427 | 77.3 (12.4-100.5) | 74.4 (6.2-101.1) | 0.874 |
| RFS: RMST [months] | 100.5±0.0 | 87.30±7.00 | 0.059* | 100.5±0.0 | 87.30±7.00 | 0.059* |
| RFS: RMST ratio | 1.15±0.10 |  | 0.079* | 1.15±0.10 |  | 0.079* |

KD: Ketogenic diet; OS: Overall survival; PFS: Progression-free survival; RFS: Relapse-free survival; RMST: Restricted mean survival time as defined by Equation (3); SD: Standard diet. \*p<0.10

Figures 1 displays the Kaplan-Meier curves for the PP population for all three outcomes. While the KD groups exhibited prolonged OS, PFS and RFS, the survival curve difference to the SD group was not statistically significant. However, when we matched the seven patients from the KD group who finished the stud regularly to seven patients from the SD group (see Materials and Methods sections) the trend of improved survival outcomes became more apparent (Supplementary Figure 1): the p-values for survival differences between both groups were 0.084, 0.066 and 0.064 for OS, PFS and RFS, respectively.

**Figure 1.**
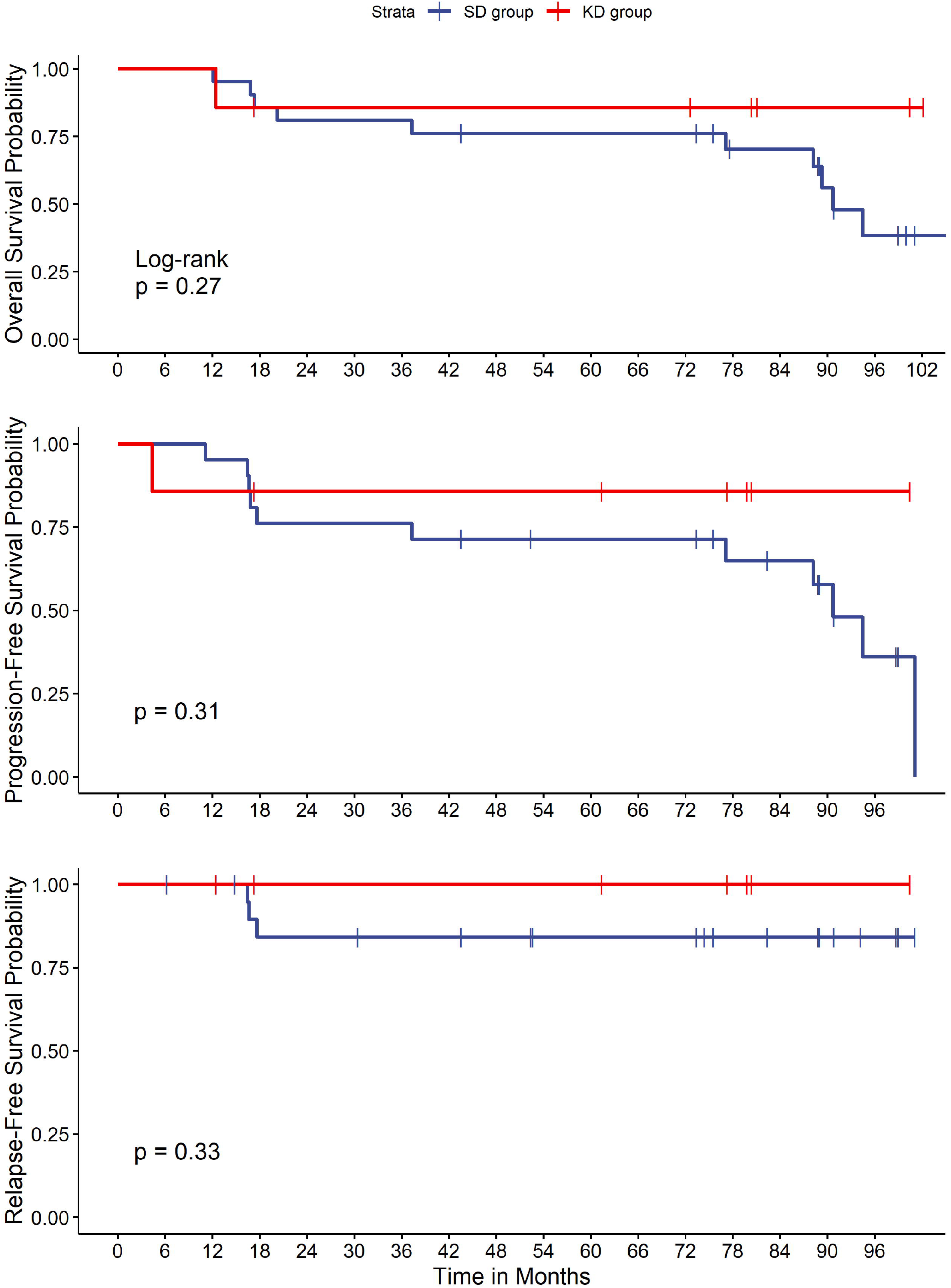
Overall, progression-free and relapse-free survival of head and neck cancer patients who finished the study regularly in the ketogenic diet (KD) and standard diet (SD) groups.

### Survival analysis for rectal cancer patients

Table 3 shows the survival statistics for the rectal cancer cohort. The length of follow-up was significantly shorter in the KD than in the SD group for OS in the ITT population. Regarding the outcomes, there were no statistically significant differences between both groups in the RMSTs. There were also no statistically significant differences in OS, PFS or RFS curves between both groups in the PP population (Figure 2) and the propensity score-matched sample (Supplementary Figure 2).

**Table 3:** Survival statistics of the intention-to-treat (ITT) and per protocol (PP) populations of rectal cancer patients.

|  | Intention-to-treat population |  |  | Per protocol population |  |  |
| --- | --- | --- | --- | --- | --- | --- |
| Parameter | KD group (N=23) | SD group (N=25) | p-value | KD group (N=18) | SD group (N=23) | p-value |
| OS: Follow-up time [months] | 62.0 (25.8-127.1) | 92.9 (16.5-122.4) | 0.023** | 61.9 (27.1-127.1) | 99.2 (16.5-122.4) | 0.065* |
| OS: RMST [months] | 96.24±8.15 | 98.48±7.28 | 0.838 | 94.82±9.32 | 99.97±7.51 | 0.667 |
| OS: RMST ratio | 0.98±0.12 |  | 0.838 | 0.95±0.12 |  | 0.669 |
| PFS: Follow-up time [months] | 59.0 (14.0-121.0) | 62.0 (3.2-113.3) | 0.703 | 60.4 (14.1-121.0) | 62.0 (7.1-113.3) | 0.922 |
| PFS: RMST [months] | 91.83±9.29 | 75.26±8.77 | 0.195 | 90.68±10.48 | 76.90±8.84 | 0.315 |
| PFS: RMST ratio | 1.22±0.19 |  | 0.198 | 1.18±0.20 |  | 0.312 |
| RFS: Follow-up time [months] | 59.0 (6.1-121.0) | 65.1 (15.5-112.2) | 0.256 | 60.4 (14.1-121.0) | 65.1 (15.5-112.2) | 0.453 |
| RFS: RMST [months] | 99.20±6.98 | 92.23±7.21 | 0.487 | 96.24±8.41 | 90.81±7.79 | 0.633 |
| RFS: RMST ratio | 1.076±0.11 |  | 0.488 | 1.060±0.13 |  | 0.633 |

KD: Ketogenic diet; OS: Overall survival; PFS: Progression-free survival; RFS: Relapse-free survival; RMST: Restricted mean survival time as defined by Equation (3); SD: Standard diet. \*p<0.10; \*\*p<0.05

**Figure 2.**
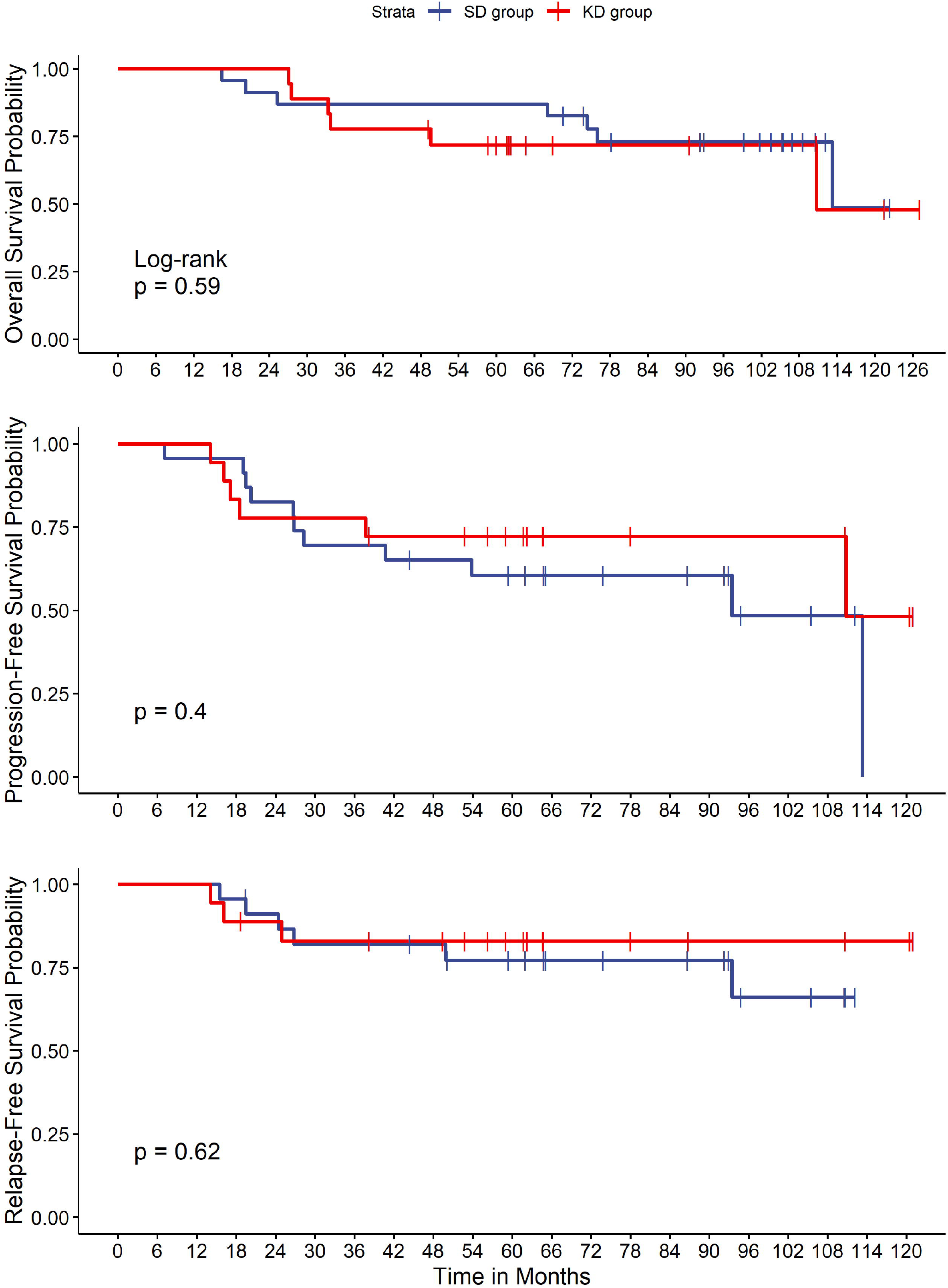
Overall, progression-free and relapse-free survival of rectal cancer patients who finished the study regularly in the ketogenic diet (KD) and standard diet (SD) groups.

It is noteworthy that within the PP population, three male patients from the KD group had refused to undergo surgery; all of them developed disease progression (two locoregional relapses) and ultimately died. In contrast, only one patient from the SD group did not undergo surgery; he also developed locoregional disease progression but was still alive at the time of last follow-up (70.6 months).

### Pooled survival statistics

In order to increase statistical power, we simulated two randomized controlled trials with the head and neck and rectal cancer patient data by matching patients from the KD group with patients from the SD group as described in the Materials and Methods section. The PTV was among the covariates when matching the rectal cancer patients, because both groups differed significantly with respect to this covariate as shown in Table 1. After matching, there were no significant differences in any of the variables displayed in Table 1.

After matching the rectal and HNC patient cohorts, both datasets were pooled and survival analysis was conducted on this pooled dataset. The numerical results are shown in Table 4, while Figure 3 displays the corresponding Kaplan-Meier curves. While OS was not significantly longer in the KD group, there was a trend for a longer restricted mean RFS time and a significantly prolonged PFS time in the KD group compared to the SD group. The difference in PFS curves almost reached the threshold of statistical significance in the log rank test (Figure 3; p=0.054).

**Table 4:** Survival statistics of the pooled patient dataset consisting of rectal and head and neck cancer patients with an equal number of treatment and control group patients and a balanced distribution of covariates.

|  | **Pooled dataset** |  |  |
| --- | --- | --- | --- |
| **Parameter** | **KD group (N=25)** | **SD group (N=25)** | **p-value** |
| OS: Follow-up time [months] | 62.3 (12.4-127.1) | 78.2 (12.1-121.5) | 0.304 |
| OS: RMST [months] | 96.09±8.09 | 86.33±8.45 | 0.404 |
| OS: RMST ratio | 1.11±0.15 |  | 0.404 |
| PFS: Follow-up time [months] | 61.7 (4.3-121.0) | 59.4 (7.1-113.3) | 0.803 |
| PFS: RMST [months] | 93.33±8.92 | 65.61±8.75 | 0.027** |
| PFS: RMST ratio | 1.42±0.24 |  | 0.032** |
| RFS: Follow-up time [months] | 61.7 (12.4-121.0) | 73.8 (6.2-112.2) | 0.778 |
| RFS: RMST [months] | 100.11±6.53 | 79.80±8.71 | 0.062* |
| RFS: RMST ratio | 1.26±0.16 |  | 0.074* |

KD: Ketogenic diet; OS: Overall survival; PFS: Progression-free survival; RFS: Relapse-free survival; RMST: Restricted mean survival time as defined by Equation (3); SD: Standard diet. \*p<0.10; \*\*p<0.05

**Figure 3.**
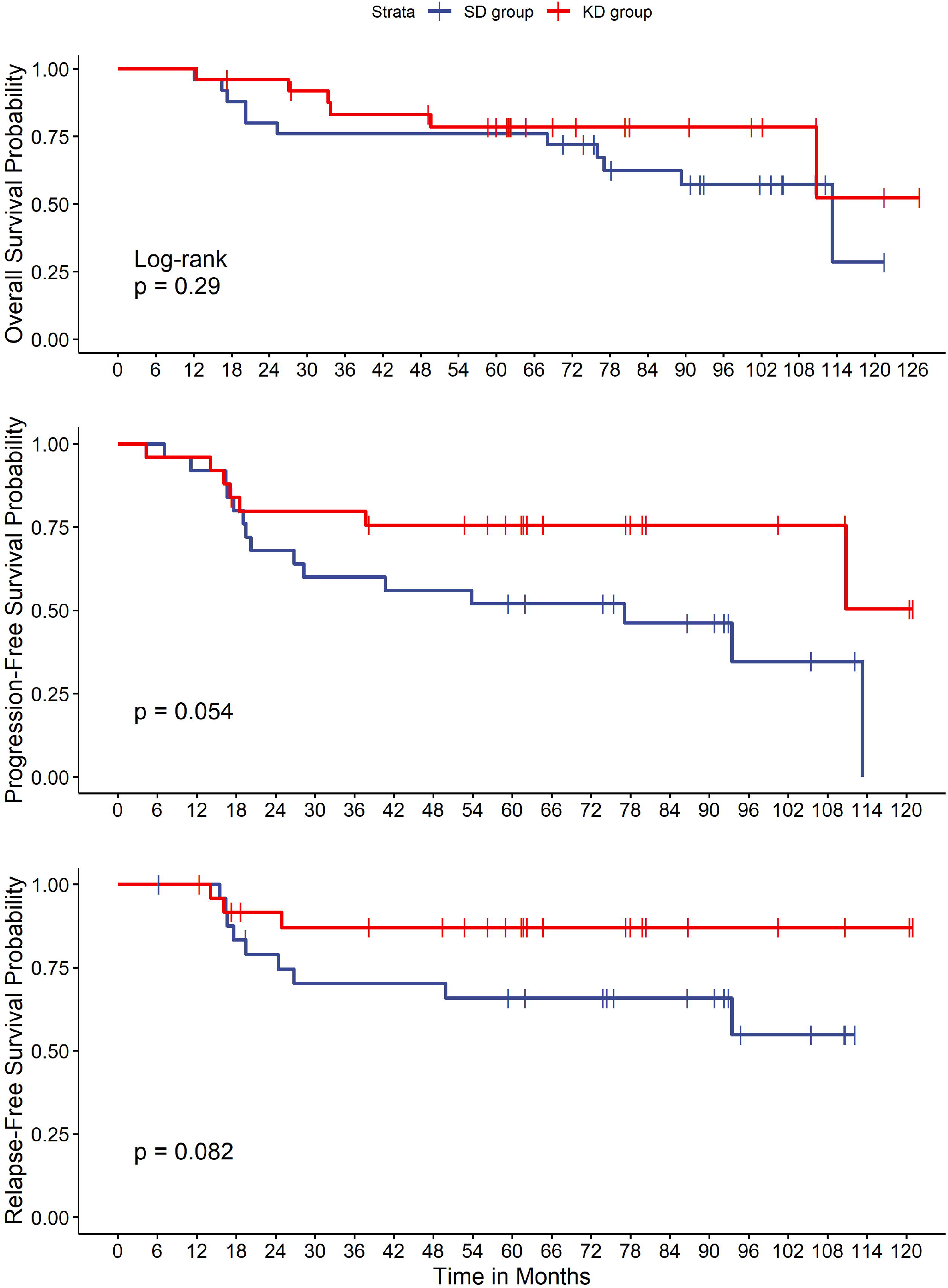
Kaplan-Meier plots for the pooled dataset. The dataset consists of both rectal and HNC patients who first had been matched using propensity score matching in order to simulate a randomized controlled trial. See Materials and Methods for details.

## Discussion

Using prospective follow-up data from our KETOCOMP trial, we here show that HNC or rectal cancer patients consuming a KD in general had better survival outcomes than control patients did on their SD. While the differences between both groups were not statistically significant when analyzed separately for each tumor entity, statistical power was increased by pooling both cohorts together, which yielded a significantly longer PFS in the KD group (Table 4). In addition, there was a trend was longer RFS in the HNC cohort as well as the pooled dataset.

According to theoretical considerations, a KD is expected to eventually improve clinical outcomes in HNC [20,21] and rectal cancer [22,23] patients. Schroeder et al. [24] have shown that after a few days of switching to a KD, tumor lactate levels were reduced in HNC patients. Given that tumor lactate levels exert a radio-protective effect in HNC tumors [25], our result of improved locoregional tumor control in the HNC cohort is consistent with these findings. In the propensity score-matched cohort, there was also a trend for improved OS and PFS (Supplementary Figure S1). However, the rectal cancer cohort did not display any differences in OS, PFS or RFS times. This is despite the fact that pathological responses after neoadjuvant radio-chemotherapy were better in the KD group at the time of surgery [11]. Trials on neoadjuvant breast cancer therapy have shown that pathological complete response is only associated with disease-free survival or OS on the individual patient level, but not on a clinical trial level [26,27]. One explanation for these as well as our findings is that pathological response of the primary tumor does not reflect the therapeutic effects on micrometastases. Furthermore, after surgery had been performed, the odds of developing locoregional recurrence would have been equal between both groups regardless of the response to neoadjuvant therapy. This is because the great majority of patients discontinued the KD after study termination, so that any putative anti-tumor mechanism of the KD could no longer operate. The larger number of patients receiving no surgery after radio-chemotherapy in the KD group (3 versus 1) could further have contributed to a distortion of the survival statistics to the disadvantage of the intervention group. Finally, some clinical data indicate that OS of cancer patients consuming a KD is positively associated with the duration of the KD [28,29], and the KD duration in our patients was relatively short compared to the follow-up time, which could also be a reason why we did not observe statistically significant differences in OS.

A meta-analysis of mouse studies has shown that among 15 cancer types, pancreatic cancer, glioblastoma, stomach cancer and HNC had the highest evidence for synergistic anti-tumor effects of a KD combined with oncological therapies [8]. While evidential support for anti-tumor effects of a KD in colon cancer was also strong when used as a monotherapy, the evidence for synergistic effects was not significant. Assuming biological similarity between colon and rectal cancer, the results of this preclinical meta-analysis would be consistent with our findings that HNC patients responded better to synergistic treatment in terms of PFS improvements than rectal cancer patients. Only a few other studies investigated the application of a KD during treatment of head and neck or rectal cancer patients. Ma et al. conducted a phase I clinical trial in patients with locally advanced HNC undergoing radio-chemotherapy with or without a KD [21]. İyikesici presented preliminary results of combining a KD with metabolically supported chemotherapy, hyperthermia and hyperbaric oxygen therapy in stage II-IV rectal cancer patients [30]; while he concluded that the survival data were promising for this combinatorial protocol, a control group not eating a KD was lacking, so that no conclusions can be drawn regarding putative anti-cancer effects of the KD. A retrospective analysis from Detroit, USA, found that rectal cancer patients consuming ≥40% energy from fat and <100g carbohydrates/day had an approximately 50% reduced risk of cancer-specific death after radiotherapy, a result that approached the threshold for statistical significance (HR 0.49, 95% CI 0.23-1.02) [31].

Findings from clinical trials investigating the KD as a complementary treatment in other tumor entities are mixed. A recently published phase II trial found evidence for synergistic effects of a KD combined with chemotherapy in pancreatic cancer patients, with improved PFS and OS in the KD group [32]. A study conducted in Tehran, Iran, found that a KD in addition to neoadjuvant chemotherapy significantly prolonged survival of breast cancer patients, but no such effect was found for metastasized patients receiving chemotherapy [33]. The majority of clinical trials reporting survival outcomes focused on patients with high-grade gliomas, but their findings are limited, either because a control group was lacking [34–36] or survival outcomes were not statistically different between KD and control groups [37]. Therefore, our study adds important data to the field of KD and cancer research.

A few limitations of our study have to be pointed out, however. Firstly, the sample sizes of the rectal cancer and HNC cohorts were rather small, which could have diminished the ability to discover statistically significant effects. The KETOCOMP study was performed in a community hospital without financial support from industry or other sources, and we believe that larger institutions would be better suited to perform larger trials in the future. Secondly, the great majority of our patients quit the KD after study termination, which could have negatively impacted survival statistics. We also did not track or control for other lifestyle factors such as physical exercise that could have influenced the course of the disease after radiotherapy. Thirdly, the degree of ketosis and the ratio between glucose and ketone body concentrations, which is proposed as a surrogate measure to track the effects of ketogenic metabolic therapy [6], were also not accounted for in this analysis.

The strength of this study, however, is that patients were prospectively followed for long periods that exceeded five years for most patients without clinical events of interest. It is therefore one the clinical trials with the longest follow-up of cancer patients who used a KD in conjunction to their standard-of-care therapy. Its results, indicating the potential of anti-tumor effects of KDs and radiotherapy in select patient cohorts could guide the design of future trials and could be used in future meta-analyses.

In conclusion, our analysis reveals weak evidence for the hypothesis, that a KD added to radiotherapy improves PFS and/or RFS, but without a significant effect on OS. Because a detrimental effect of the KD on any of the three survival outcomes was not observed, our study adds to the growing literature confirming the safety of ketogenic metabolic therapy.

## Data Availability

Research data are stored in an institutional repository and will be shared upon request to the corresponding author.

