## Supplementary Material for "Survival outcomes of rectal and head and neck cancer patients receiving radio-(chemo)therapy with a ketogenic diet. Results from a controlled clinical study (KETOCOMP)"

**Supplementary Table 1: Baseline characteristics of the intervention and control groups in the propensity score-matched datasets**

| | **Head and neck cancer** | | | **Rectal cancer** | | |
|---|---|---|---|---|---|---|
| **Parameter** | **KD group (N=7)** | **SD group (N=7)** | p-**value** | **KD group (N=18)** | **SD group (N=18)** | p-**value** |
| Gender | | | 0.592 | | | 1 |
| *Male* | 5 (71%) | 3 (43%) | | 12 (67%) | 11 (61%) | |
| *Female* | 2 (29%) | 4 (57%) | | 6 (33%) | 7 (39%) | |
| Age [years] | 65 (61-75) | 65 (60-72) | 0.796 | 56 (38-77) | 61 (43-76) | 0.254 |
| Karnofsky Index | | | 1 | | | 0.078 |
| 70 | 4 (57%) | 4 (57%) | | 8 (47%) | 2 (12%) | |
| 80 | 2 (29%) | 3 (43%) | | 6 (35%) | 8 (47%) | |
| 90 | 1 (14%) | 0 (19.0%) | | 3 (18%) | 7 (41%) | |
| T stage | | | 0.449 | | | 0.692 |
| 1 | 0 | 1 (14%) | | 0 | 0 | |
| 2 | 5 (71%) | 2 (29%) | | 1 (6%) | 3 (17%) | |
| 3 | 1 (14%) | 2 (29%) | | 15 (83%) | 14 (78%) | |
| 4 | 1 (14%) | 2 (29%) | | 2 (11%) | 1 (6%) | |

| | | | | | | |
|---|---|---|---|---|---|---|
| N stage<br>0<br>1<br>2<br>3<br>+<br>X | <br>2 (29%)<br>1 (14%)<br>4 (57%)<br>0<br>0<br>0 | <br>3 (43%)<br>1 (14%)<br>2 (29%)<br>1 (14%)<br>0<br>0 | 0.767 | <br>6 (33%)<br>7 (39%)<br>2 (11%)<br>0<br>3 (18%)<br>0 | <br>4 (22%)<br>9 (50%)<br>1 (6%)<br>0<br>3 (17%)<br>1 (6%) | 0.851 |
| Body weight [kg] | 64.2 (55.2-76.7) | 60.9 (53.9-79.9) | 0.522 | 80.8 (55.9-105.7) | 78.9 (52.1-111.8) | 0.743 |
| BMI [kg/m$^2$] | 22.5 (19.3-26.1) | 23.8 (17.8-27.3) | 0.805 | 26.8 (20.2-35.0) | 26.4 (19.5-32.8) | 0.606 |
| Phase angle [°] | 4.33 (3.98-4.74) | 4.1 (4.04-4.56) | 0.223 | 4.94 (3.74-6.59) $ | 5.06 (4.05-5.97) | 0.520 |
| Diabetes<br>*No*<br>*Yes* | <br>5 (71%)<br>2 (29%) | <br>7 (100%)<br>0 (0%) | 0.390 | <br>16 (89.0%)<br>2 (11.0%) | <br>14 (78%)<br>4 (22%) | 0.658 |
| Smoking status<br>*No*<br>*Active*<br>*Formerly* | <br>1 (14%)<br>0<br>6 (86%) | <br>1 (14%)<br>3 (43%)<br>3 (43%) | 0.192 | <br>12 (67%)<br>0 (8.7%)<br>6 (33%) | <br>11 (61%)<br>2 (11%)<br>5 (28%) | 0.558 |

| | | | | | | |
|---|---|---|---|---|---|---|
| Tobacco consumption [packyears] | 17.5 (0-60) | 40 (0-55) | 0.830 | 0 (0-30) | 0 (0-35)[$] | 0.891 |
| Systemic therapy<br>*No*<br>*Yes* | <br>3 (43%)<br>4 (57%) | <br>3 (43%)<br>4 (57%) | 1 | <br>0<br>18 (100%) | <br>1 (6%)<br>17 (94%) | 1 |
| Radiation dose [Gy] | 58 (50-71) | 60 (50.4-75) | 0.369 | 50.2 (50-61.2) | 50 (50-55.4) | 0.149 |
| Radiation therapy fractions | 30 (25-34) | 30 (28-36) | 0.476 | 26 (25-31) | 25 (25-30) | 0.124 |
| PTV [$cm^3$] | 636 (155-1278) | 707 (471-1359) | 0.805 | 1254 (944-1845) | 1376 (950-1838) | 0.161 |

Continuous and categorical variables are presented as median (range) and counts (frequencies), respectively. BMI: body mass index; KD: ketogenic diet; PTV: Planning target volume; SD: standard diet. [$] Data missing for one patient.

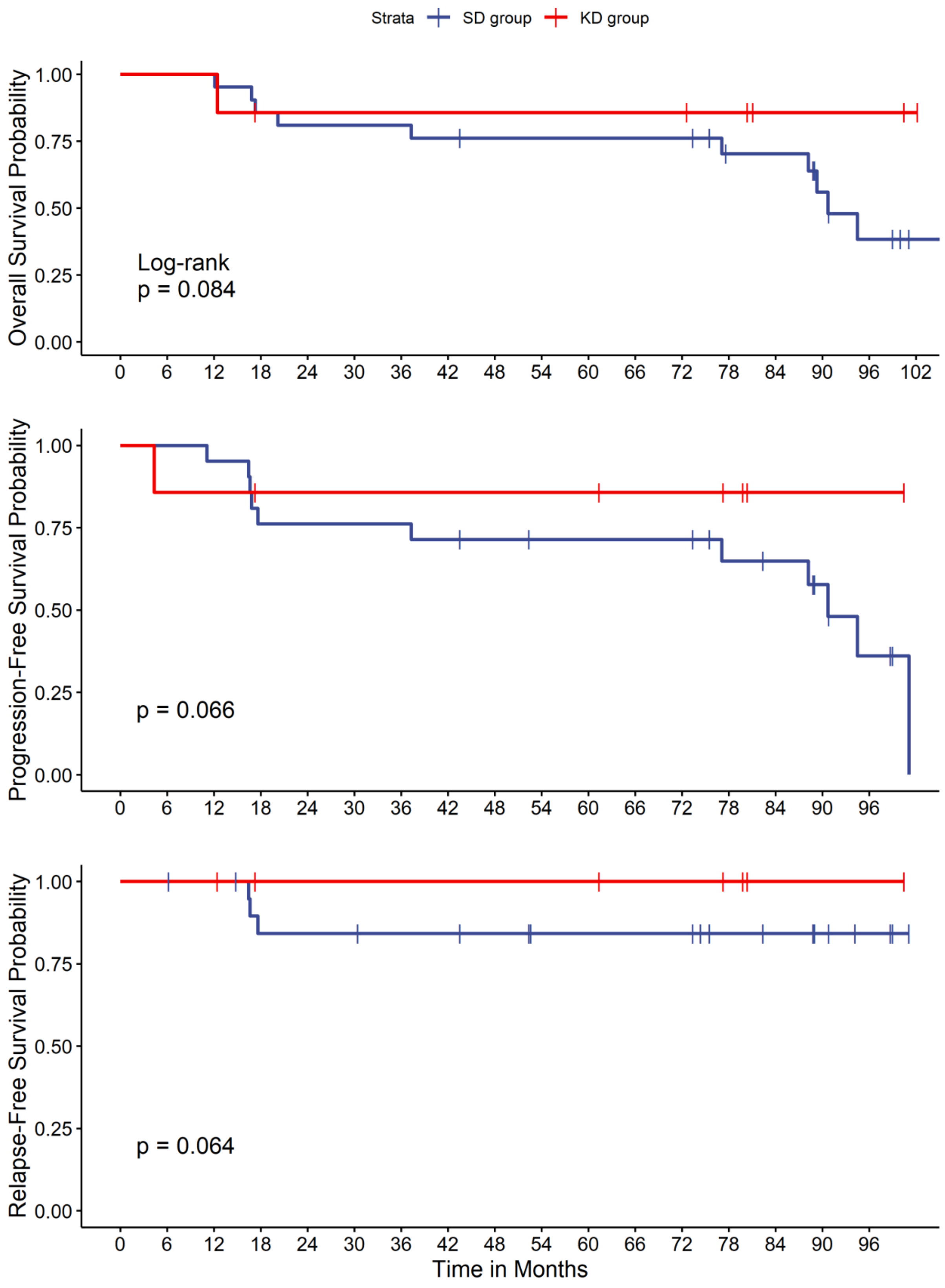


Supplementary Figure 1: Overall, progression-free and relapse-free survival in the propensity score-matched sample of head and neck cancer patients.

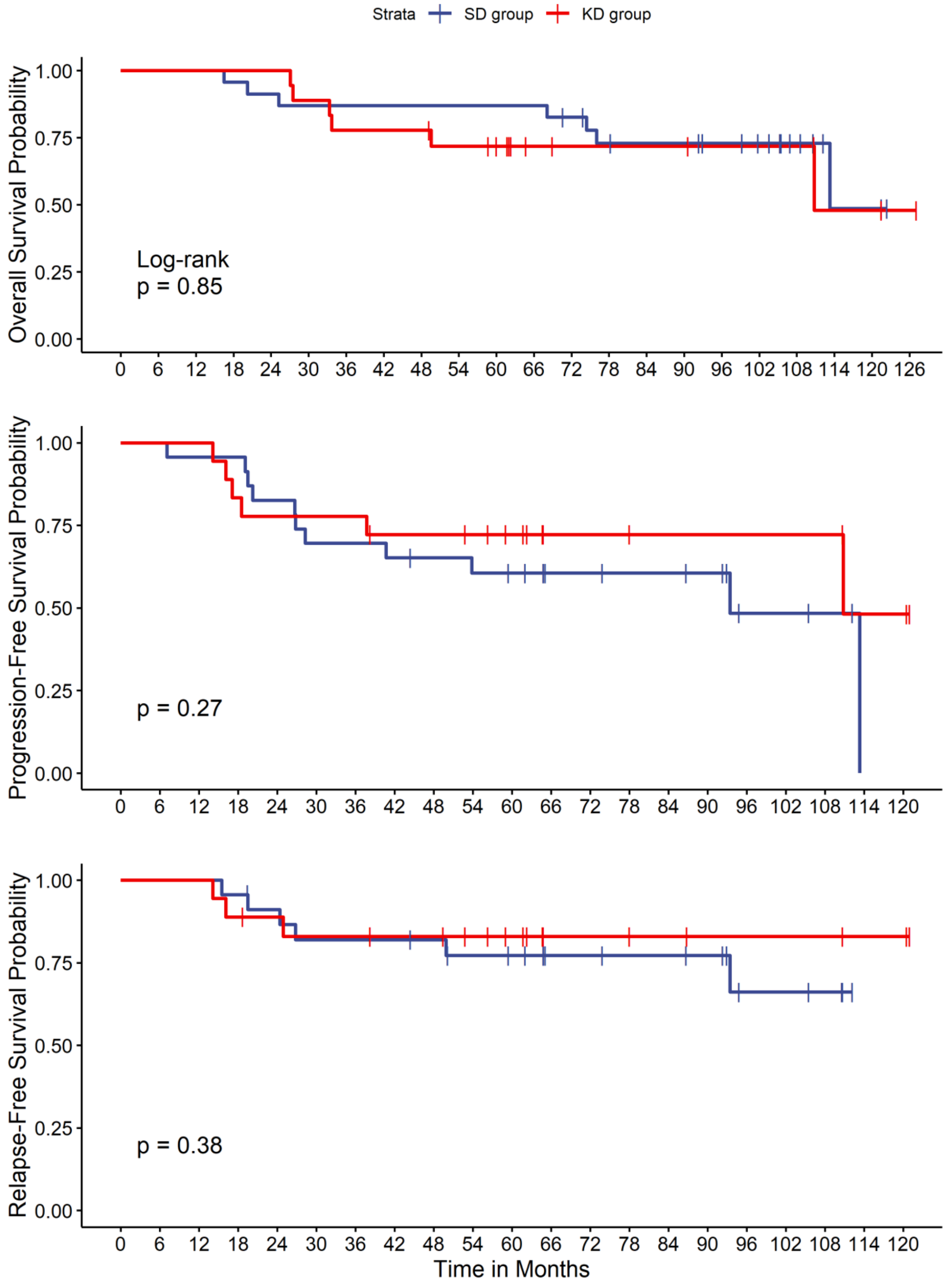


Supplementary Figure 2: Overall, progression-free and relapse-free survival in the propensity score-matched sample of rectal cancer patients.
